# Human trafficking and sexual exploitation at U.S. colleges/universities: Policy implications to halt it

**DOI:** 10.64898/2025.12.31.25342184

**Authors:** L.A. Urada, A. Weitensteiner

## Abstract

**Objectives:** Human trafficking is a priority concern in the U.S, but few research studies have investigated the nature and extent of its occurrence on U.S. college campuses. The objective of this research was to identify the nature and extent of human trafficking/sexual exploitation among college students in San Diego County and Imperial Valley, California, where some of the highest documented rates of human trafficking occur nationally.

**Methods:** College students (n = 971) from 12 campuses in Southern California responded to fliers posted on their campuses by completing online self-administered surveys.

**Results:** Nearly one in five students (18%) reported experiencing human trafficking in college; 13% reported having heard, seen, or suspected trafficking recruitment occurring on campus. Further, students reported having known someone who sold sex or was trafficked as a college student (42%), while also describing the nature of their peers’ sexual exchanges as having been forced (12%), threatened (12%), coerced (26%), or deceived into selling sex (24%). They experienced physical abuse/violence (51%); rape, emotional/psychological harm (43%), and sexual assault/violence (42%) in commercial sex.

**Conclusions:** Strategies to prevent human trafficking recruitment and to protect college students may need to mandate content for college student orientations in their first year and to test and evaluate these strategies to curb this public health crisis. Recommendations are: 1) expand Title IX implementation to encompass human trafficking/commercial sexual exploitation information at all universities, and 2) amend or introduce state laws mandating human trafficking training at schools to include colleges/universities.

## Introduction

Human trafficking is a priority concern in the U.S. [1], yet few research studies have investigated the nature and extent of its occurrence on U.S. college campuses. The U.S. defines human trafficking as “force, fraud, or coercion” into commercial sex (or other services). Anyone under age 18 engaged in commercial sex is considered trafficked because they cannot legally consent to it [2]. Since its landmark legislation, the Victims of Trafficking and Violence Protection Act of 2000, guides nations’ scores on how well nations address human trafficking [2]. However, 25 years later, after multiple iterations, legal definitions confer automatic human trafficking status only on youth engaged in commercial sex under age 18. However, Transitional Age Youth over age 18 face similar conditions and risks for exploitation during their transition to adulthood. Science tells us the executive functioning of human brains continues to develop through age 25 or even 30 [3].

Furthermore, the transition into college often involves stepping away from familiar social networks and seeking new connections, which can leave students vulnerable to manipulation. Financial instability is also typical as many students take out loans and incur debt to attend school, further increasing their susceptibility, and in many cases, leading to homelessness (e.g., living out of their cars) [4]. Additionally, traffickers may take advantage of the culture of substance use experimentation that exists on many campuses to exploit and exert control over students. The lure of alternative ways to make money and support themselves, especially those with little family support, makes college-age students even more vulnerable to sexual exploitation.

Therefore, the objective of this research was to identify the nature and extent of human trafficking and sexual exploitation occurring among college students in two southern California regions: San Diego County and Imperial Valley, U.S.-Mexico border regions with some of the highest documented rates of human trafficking nationally [5].

## Methods

### Design and setting

We applied a cross-sectional self-administered survey design to examine experiences of human trafficking and sexual exploitation among college students in southern California. Students were recruited from 12 different universities and colleges, including four-year public and private universities and community colleges within San Diego and Imperial Counties in California.

### Ethical approach

Study approval was obtained from the institutional review board of San Diego State University (IRB#: HS-2022-0162). Study approval was also obtained from the eleven other colleges and universities in San Diego and Imperial counties within southern California from 2022 to 2023. Informed consent was obtained after participants were made aware of the study purpose, design, and anonymous nature of the survey. The voluntary nature of participation was emphasized, and at several points in the online survey participants were prompted with a reminder that they could withdraw (exit) the survey at any time. Participants who selected that they consented to the study were then taken to a page to start the survey. A resource list including mental health and human trafficking specific services, located both on and off campus, and tailored to each college or university was provided at beginning and end of the survey. Participants who declined to consent were also provided with this list of resources. A prompt within the survey provided students the option of requesting researchers contact them to speak with someone (such as a licensed professional or human trafficking survivor) and to provide a direct connection to human trafficking and or mental health resources. Survey responses were anonymous. Participants’ contact information to receive remuneration for their participation in the study was collected via a separate linked form that was not connected to their survey data.

### Recruitment

Students were recruited via targeted placement of study flyers on college campuses in areas frequented by students and approved by the school and/or IRB, giving students an equal chance to respond to the flyer. These areas included bathroom stalls, campus bulletin boards, hallways, student centers, and libraries. Flyers described the study and included a QR code and survey link that directed students to the online survey, hosted on Qualtrics survey platform. Informed by an advisory board of human trafficking survivor leaders, recruitment fliers intentionally avoided terms like “human trafficking” or “modern slavery.” Students may not immediately identify their experiences as human trafficking. Therefore, the term “sexual exploitation” was used on the flyer, asking for those to participate if they experienced or knew someone who experienced sexual exploitation as a college student.

To be eligible to participate in the study, participants must have been 1) 18 years or older, and 2) currently enrolled students or alumni of one of the participating institutions. Participants completed the survey on their own electronic device at a time and place of their choosing. Surveys took approximately 15 to 45 minutes to complete. Participants received a 15-dollar Amazon gift card for their time and effort.

### Study development and measures

The survey instrument was developed in collaboration with a human trafficking survivor leader advisory group. Our main outcome measure of human trafficking was defined as experiencing force, fraud, or coercion into exchanging sex for money or something of monetary or other value, aligning with U.S. federal definitions [2]. We made a distinction in the survey between human trafficking and exchanging sex for something of monetary or other value free of coercion, force, or fraud. The survey asked about both separately.

Surveys included questions around sexual exploitation, labor and sex trafficking, attitudes related to these acts, motives for selling sex, and gender norms. These questions were informed by previous research and consultation with human trafficking survivors [6,7] and surveys with university students mostly outside of the U.S. Additional domains included sociodemographic characteristics, substance use, stigma, and mental health. Most items were multiple choice. Some questions included Likert type scales or open-ended responses to allow for students to elaborate on their responses. Full surveys were further reviewed and vetted by individuals with lived experience of commercial sex exchanges and human trafficking.

### Analysis

Stata statistical software package was used for cleaning and analysis of data. Descriptive statistics, including frequencies and percentages, were used to summarize demographics and experiences of trafficking and sexual exploitation among the survey respondents. Stata statistical software package was used for cleaning and analysis of data.

## Results

College students (n = 971) from 12 campuses in Southern California, primarily located in the San Diego and Imperial Valley regions, completed the surveys. However, students also described multiple affiliations, with some having spent time at Los Angeles County colleges/universities, for example. Participants were predominantly undergraduate, female (69%), male (25%), and gender diverse individuals (6%). Two-thirds (66%) reported their sexual orientation as heterosexual; 14% identified as bisexual, 7% as gay/lesbian, and 9% as other. They were 38% White, 36% Latino/a, 18% Asian/Pacific Islander, 17% Black, 5% American Indian/Native Alaskan, and 2% “other.” Sixty-one were former foster youth [Table 1].

**Table 1.** Demographic and sexual exploitation-related variables among college students surveyed across 12 universities in southern California (n=971)

|  | n | % |
| --- | --- | --- |
| **Demographics** |  |  |
| Gender |  |  |
| Woman | 669 | 69 |
| Man | 242 | 25 |
| Gender diverse | 55 | 6 |
| Sexual orientation |  |  |
| Heterosexual | 663 | 66 |
| Homosexual | 75 | 7 |
| Bisexual | 146 | 14 |
| Other/Questioning | 87 | 9 |
| Race and ethnicity |  |  |
| White | 373 | 38 |
| Latino/a | 348 | 36 |
| Asian/Pacific Islander | 176 | 18 |
| Black | 166 | 17 |
| American Indian/Native Alaskan | 46 | 5 |
| Other | 22 | 2 |
| Current or former foster youth | 61 | 6 |
| **Human trafficking/sexual exploitation experiences** |  |  |
| Experienced trafficking as a student | 172 | 18 |
| Knew a student who sold sex or was trafficked | 363 | 42 |
| Reported nature of sexual exchange |  |  |
| Forced into selling sex | 56 | 12 |
| Threatened into selling sex | 59 | 12 |
| Coerced into selling sex | 124 | 26 |
| Deceived or defrauded into selling sex | 112 | 24 |
| Heard, saw, or suspected trafficking recruitment on campus | 107 | 13 |
| Selling sex as a college student |  |  |
| Considered selling sex | 168 | 20 |
| Sold sex one time | 84 | 10 |
| Sold sex more than once | 94 | 11 |
| Never considered selling sex | 469 | 55 |
| Sold sex across the international border | 135 | 16 |
| Experiences arising from exchanging sex for something on monetary or |  |  |
| other value* |  |  |
| Experienced emotional or psychological harm | 268 | 43 |
| Experienced physical abuse or violence | 182 | 51 |
| Rape, sexual assault, or sexual violence | 154 | 42 |
| Told or asked to recruit others | 104 | 29 |
| Felt pressure to engage in sexual activity as a student | 335 | 38 |
| Other activities students felt pressured by someone to do |  |  |
| Transport or sell drugs | 46 | 17 |
| Perform labor | 33 | 12 |
| Exchange sex for help with schoolwork or grades | 47 | 18 |

\*the denominator is 626 for those who exchanged sex for something of monetary or other value

Nearly one in five students (18%) who answered the surveys reported having experienced human trafficking (force, coercion, fraud into exchanging sex for something of monetary or other value) during college; 13% heard, seen, or suspected trafficking recruitment occurring on campus. Further, many students reported having known someone who sold sex or was trafficked as a college student (42%), and described the nature of their peers’ sexual exchanges as having been forced (12%), threatened (12%), coerced (26%), or deceived into commercial sex exchanges (24%). Students also reported traveling across the border to sell sex or knowing students who did as college students (16%). They reported how commercial sex involvement impacted student safety and well-being. Over half experienced physical abuse or violence as a result (51%). Others experienced emotional or psychological harm (43%), rape, sexual assault, or other sexual violence (42%), and being told or asked to recruit others to sell sex (29%).

Many students described feeling pressure to engage in sexual activities (38%). They also felt pressured by someone they met on campus to transport or sell drugs (17%), perform labor (12%), or exchange sex for help with schoolwork or obtaining better grades (18%). Students described pressure as originating from another student (44%), romantic partners (23%), professors (15%), and other individuals (9%). Students also described their intentions to sell sex, with 20% having considered it, 21% selling sex at least once, and 55% never considering it.

## Discussion

The immense profitability of trafficking leads to continuously adaptive methods to lure victims. College campuses are no exception. In this study, we found that sexual exploitation and sex trafficking occurred across a range of university types: four-year public and private universities and community college districts. Research, including this one, suggests that traffickers have identified college campuses and other places where students congregate (e.g., parties, malls, clubs) as prime geographical recruitment targets [7].

The study’s finding that one in five college students responding to the self-administered surveys experienced sexual exploitation as U.S. college students on and around U.S. campuses is alarming. However, the study’s results should be interpreted with caution because not all students were surveyed as it was not a randomized sample nor a survey of all students. However, as in most surveys, student self-selected to take it and the mere fact that so many did, demonstrates that sexual exploitation and human trafficking are happening among college students, contrary to what the public may suspect. With very few, if any, studies surveying U.S. college students about their sexual exploitation, this study shed light onto the nature and extent it is happening at all.

### Policy Implications

In 2017, California enacted the Human Trafficking Prevention Education and Training Act to combat trafficking in middle and high schools through educational campaigns and training for students and teachers [8]; other states followed suit. However, experts now understand that college students, too, are vulnerable to such predators. The U.S. Department of Homeland Security’s awareness campaign, Blue Campaign, recently began distributing guidance to campus police about human trafficking only after college personnel alerted them about the campus crime [9].

Under U.S. legislation, Title IX, colleges and universities address sexual assault on campuses [10]. Under Title IX, human trafficking or commercial sexual exploitation falls under the broader categories of sexual violence, sexual harassment, and sexual assault, and are not uniformly addressed across campuses. Therefore, the state of Virginia recently enacted legislation mandating human trafficking information in first-year orientations [11]. However, formal evaluations are lacking on the effectiveness of such campaigns for college students. More research on the risk factors leading to commercial sexual exploitation among college students, and the nuances that lead to students’ exit from exploitation, are needed.

Recommendations to curb this sexual exploitation crisis include expanding 1) Title IX implementation to encompass human trafficking/sexual exploitation information at colleges/universities, particularly in new student orientations to their universities during their first year and 2) state laws mandating human trafficking training at middle/high schools to include colleges/universities. Strategies to prevent human trafficking among college students could target incoming student orientations throughout their first year. Training of college personnel (e.g., counselors, student affairs officers) could also help detect and assist students experiencing sexual exploitation, including those living in dorms away from their parents/caregivers.

## Data Availability

All data produced in the present study are available upon reasonable request to the authors.

https://socialwork.sdsu.edu/people/faculty/urada-lianne

## Acknowledgements

We are deeply grateful to the Alexander and Eva Nemeth Foundation and their Deputy Director Alicia DeLeon-Torres for providing funding and support for this project; survivor leader advisory boards, advocates, and persons with lived experience; Ebony Jones, Jessica Kim, and Marjorie Saylor; Bruna Mori and Maja Blazejewska (consultants); Drs. Arduizur Richie-Zavaleta and Ami Carpenter (other university partners); research assistants Lauren Azar-Miller who co-designed the first survey conducted initially at one university as a pilot, Derrick Ruiz, John Kennon, Christian Cacho, Kim Kritzer, Julianne Eggold, Talin Gharibian, and Ashley Weitensteiner (doctoral student and co-author of this manuscript); and especially to the college students who participated in this research and were very much impacted by sexual exploitation occurring at their campuses.

